# Data-Driven Insights on Opioid Use and Health Behavior Trends Following Decriminalization: Zero-Shot Sentiment and Behavior Analysis

**DOI:** 10.1101/2025.02.09.25321976

**Authors:** Sepehr Harfi Moridani, Chan Pang Yang, Mohammad Noaeen, Zahra Shakeri

## Abstract

Opioid decriminalization has taken on renewed urgency in regions grappling with high mortality and health-care costs. Traditional assessments often focus on legal or epidemiological data, leaving gaps in understanding how the public actually perceives and reacts to such policies. This paper introduces an AI-driven approach that applies Mistral, a Large Language Model (LLM), to a corpus of over 22,000 Reddit comments discussing British Columbia’s decriminalization policy. Our method uses zero-shot classification to track shifts in sentiment and self-reported behaviors related to opioid use and harm reduction. The findings suggest that online conversations initially reflected optimism about reduced stigma and broader acceptance of harm reduction measures, but sentiment became more mixed as policy details and lived experiences surfaced. This pattern indicates that advanced LLM-based text analysis can yield deep insights into the evolving public narrative on health interventions, informing future policymaking and healthcare strategies.

## I. INTRODUCTION

The opioid crisis has emerged as one of Canada’s most significant public health challenges, resulting in over 45,000 deaths since 2016 and a 200% rise in annual opioid-related fatalities between 2016 and 2023 [1]. Opioids include prescription analgesics such as morphine and oxycodone as well as illicit substances like fentanyl and heroin [2], [3]. The route of administration, frequency of use, and individual tolerance levels all shape the risk of harm [4], [5]. Co-occurring mental health conditions and limited access to harm reduction services can heighten the likelihood of dependence or overdose [6]. The financial toll on the healthcare system is high, with billions spent annually on emergency interventions and care [7], [8]. In response, several regions worldwide have considered or implemented decriminalization to reduce stigma and encourage treatment [9]. Oregon initially decriminalized small amounts of hard drugs in 2023 but reversed course in September 2024, citing policy and implementation hurdles [10].

British Columbia (BC) has been hit hardest within Canada, reporting the highest rate of opioid-related deaths in 2023. On January 31, 2023, BC became the first, and currently only, province to remove criminal penalties for possession of small (under 2.5 g) amounts of certain illegal substances for adults [11]. The aim is to reduce stigma and shift individuals toward health and social services rather than penalizing them under the criminal justice system. Public opinion plays a key part in the policy’s success, as it can influence how widely the policy is accepted and whether at-risk individuals feel safer seeking help. Social media platforms, especially Reddit, provide a large pool of timely and organic user perspectives on policy changes. Unlike traditional surveys, which can be limited by recall bias or social desirability effects, Reddit discussions often include candid reflections on drug use, harm reduction, and policy outcomes [12].

This study leverages more than 22,000 Reddit comments to investigate two main research questions. First, we explore how sentiment toward people who use opioids has shifted since decriminalization, using labels ‘Negative’, ‘Neutral’, and ‘Positive’. Second, we examine whether self-reported behaviors have changed, focusing on opioid consumption patterns and mentions of harm reduction practices. Traditional text mining approaches often face challenges with colloquial expressions and context-dependent meanings. To address these issues, we employ *Mistral 7B Instruct v0*.*3*, a Large Language Model (LLM) optimized for zero-shot or prompt-based tasks. Its grouped-query attention and sliding-window mechanism enable more efficient processing of long Reddit threads, while its generative foundation supports direct text classification without extensive fine-tuning. Our method goes beyond keyword matching by assessing the context in which users discuss opioid use or describe harm reduction strategies. The resulting insights can help inform policymakers and health organizations on how the public perceives decriminalization and whether it translates into safer behaviors or ongoing controversies around opioid misuse.

## II. Methods

### A. Problem Formulation and Data Collection

We frame our analysis of sentiment and self-reported behavior within online discussions on opioid decriminalization as a text classification problem. Let 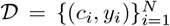 be a dataset of *N* comments, where each comment *c*_*i*_ is a sequence of tokens 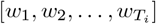, and *y*_*i*_ denotes one or more labels assigned to *c*_*i*_. We define two classification tasks:

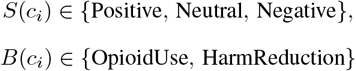

where *B*(*c*_*i*_) is actually a pair (*u*_*i*_, *h*_*i*_) indicating the presence or absence of user-reported opioid use (‘Yes’, ‘No’, ‘Not clear’) and harm reduction behaviors (‘Yes’, ‘No’, ‘Not clear’).

To construct *D*, we assembled a corpus of Reddit posts and comments containing the phrase ‘BC Decriminalization’ in content published after January 31, 2023. Each post was retrieved with metadata (title, URL, publication date), then all top-level comments and nested replies were extracted. The final dataset comprises 22,132 comments from January 2023 through October 2024. We recorded each comment’s text, number of upvotes, publication timestamp, subreddit name, and reply count. Personally identifiable information was removed to preserve user privacy. Figure 1 shows the distribution of comments over time, including a notable surge in early 2024.

**Fig. 1:**
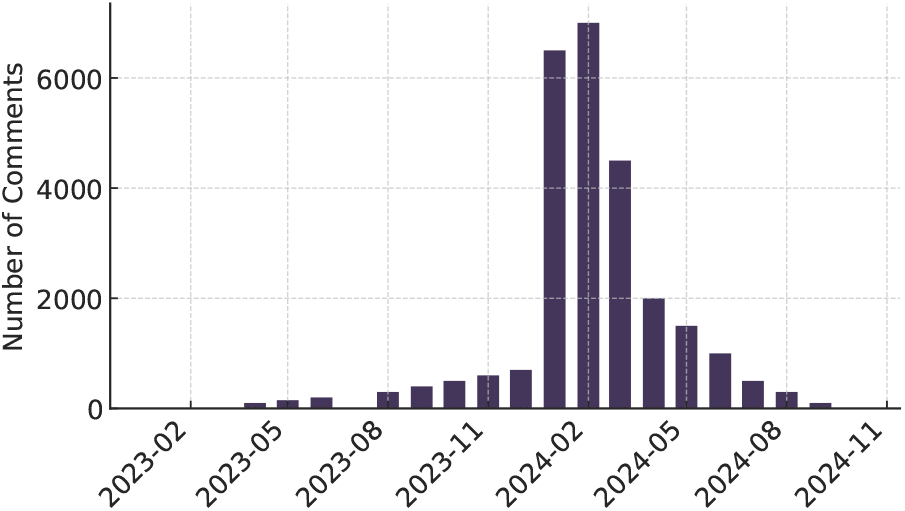
Distribution of comments over time showing a significant peak in early 2024, which may correspond to increased discussions around a key event or policy change.

### B. Large Language Model Setup

#### Model Selection

We employ a Transformer-based large language model *M*_*θ*_ to infer both *S*(*c*_*i*_) and *B*(*c*_*i*_). Figure 2 summarizes the architectural differences among several well-known Transformers:

- *GPT* [13] and *LLaMA* [14] are decoder-only, auto-regressive models that handle generation effectively but can be resource-intensive for very long sequences.
- *BERT* [15] and *RoBERTa* use encoder-only bidirectional self-attention, which excels in masked-language tasks but often requires additional heads or fine-tuning for instruction-based inference.
- *Mistral* [16] (our chosen model) introduces features such as *Grouped-Query Attention (GQA)* and a *Sliding-Window Self-Attention* mechanism. Although still decoder-based, these optimizations reduce memory overhead and improve local context processing in longer inputs.

Concretely, for a given input sequence {*x*_1_, *x*_2_, …, *x*_*L*_}, each layer produces hidden representations *H*^*l*^. In standard self-attention, head *h* computes:

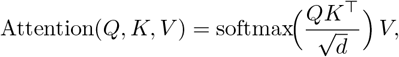

where *Q, K, V* are linear maps of *H*^*l*−1^ and *d* is the head dimension [17]. By contrast, Mistral introduces GQA (reducing redundant projection steps) and a sliding-window scheme that partitions or groups tokens for more localized attention. This is especially helpful when processing lengthy Reddit threads.

*Encoder-only* models like BERT or RoBERTa require domain-specific fine-tuning or additional classifier heads for a prompt-based approach. *GPT-like* or *LLaMA-like* decoders can handle zero-shot prompts, but at higher parameter counts or memory usage. *Mistral 7B Instruct v0*.*3* balances generative instruction-following with moderate resource requirements, crucial for analyzing thousands of comments with minimal overhead. Its GQA and sliding-window attention natively improve handling of longer inputs, matching the nature of nested Reddit discussions on opioid decriminalization.

#### Prompt Design and Zero-Shot Implementation

We adopt a zero-shot approach that supplies the model *M*_*θ*_ with carefully crafted textual prompts. One prompt requests a sentiment label (‘Positive’, ‘Neutral’, or ‘Negative’). Another prompt checks for self-reported opioid use and harm reduction references, each categorized as ‘Yes’, ‘No’, or ‘Not clear’. By appending the comment body (and optional metadata like publication date) to these instructions, we steer the model toward structured outputs (*S*(*c*_*i*_), *B*(*c*_*i*_)). This design eliminates the need for large annotated training sets or supervised fine-tuning.

**Fig. 2:**
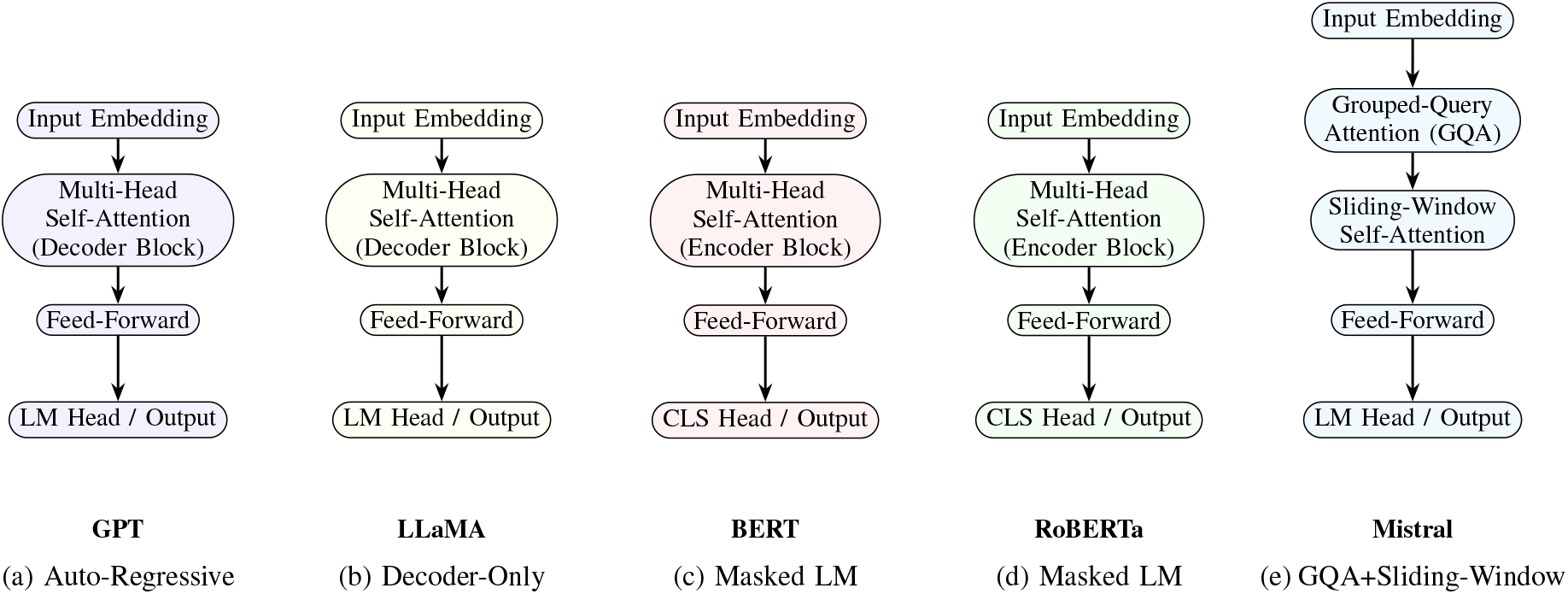
Simplified Transformer architectures for GPT, LLaMA, BERT, RoBERTa, and Mistral. GPT and LLaMA are decoder-only, auto-regressive models designed for sequential token generation. BERT and RoBERTa follow an encoder-only, masked language model (MLM) architecture for token prediction within context. Mistral introduces Grouped-Query Attention (GQA) and Sliding-Window Self-Attention, enabling efficient processing of longer texts.

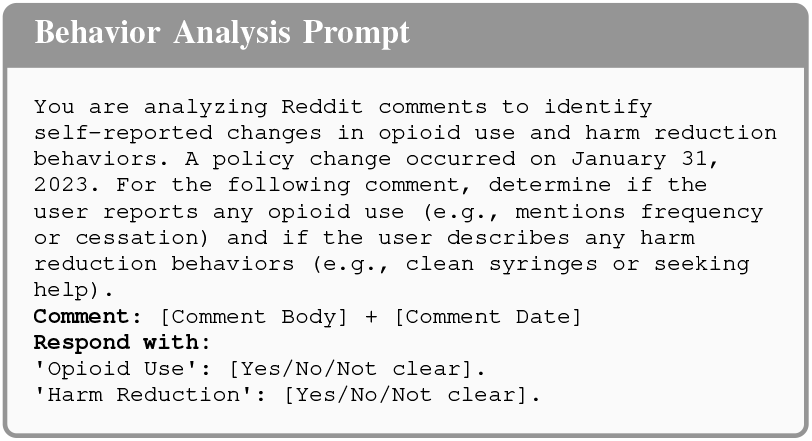

In practice, the raw model outputs are parsed to identify the final labels. This process preserves user privacy because no personally identifiable user data is retained or exposed beyond each comment’s text. By combining the sentiments *S*(*c*_*i*_) with behavioral labels *B*(*c*_*i*_), we can quantify shifts in public discourse regarding opioid use and harm reduction strategies over time, coinciding with the policy enacted on January 31, 2023.

## III. Results and Discussion

### A. Sentiment Analysis

Figure 3’s left panel shows the distribution of *S*(*c*_*i*_) categories. Negative sentiment dominates, suggesting a widespread belief that the policy might not adequately curb opioid harms or that it removes important legal deterrents. Neutral discussions are also substantial, featuring informational posts about policy details or updates on local health services. Positive sentiment is the smallest segment, reflecting a minority of comments that explicitly support decriminalization or share hopeful anecdotes about reduced stigma. This finding resonates with prior research noting varied public reactions to harm reduction policies [6].

**Fig. 3:**
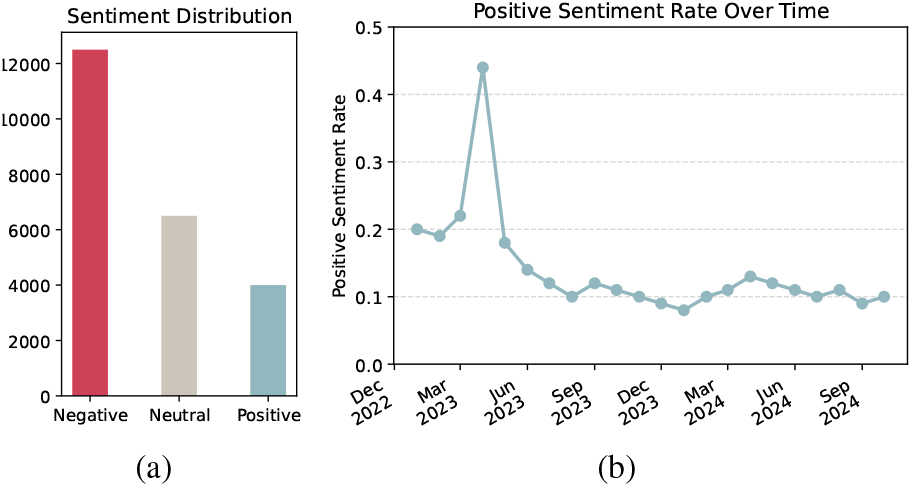
(a) Distribution of Reddit comments by sentiment (‘Negative’, ‘Neutral’, ‘Positive’). (b) Month-to-month trend in the proportion of ‘Positive’ sentiment, peaking soon after the policy launched.

Figure 3’s right panel tracks shifts in positive sentiment over time. Our Mistral-based zero-shot classification indicates an early surge in ‘Positive’ comments immediately after January 31, 2023. Some users linked this optimism to the possibility of redirecting law enforcement resources toward treatment and recovery. Others mentioned personal stories where community agencies seemed more open to providing support, reflecting the sense that criminal penalties were a primary barrier. However, the positivity fraction declines gradually as more individuals expressed skepticism. The conversation became broader, incorporating concerns about rising drug-related activities in public spaces and persistent issues of mental health support or housing instability. This complex arc highlights how short-term optimism surrounding new policies can give way to ongoing debates about feasibility and long-term outcomes.

### B. Behavioral Analysis

Figure 4 shows how our model labeled two behavioral indicators, *B*(*c*_*i*_). Specifically, it depicts the rate of ‘No Harm Reduction’ mentions (i.e., no explicit mention of safer practices) and the self-reported opioid use rate. The steady drop in no-harm mentions suggests an uptick in talk around safer practices, including references to naloxone training or supervised injection sites. Some commenters remarked that they felt more comfortable asking for help without fear of legal repercussions. These accounts align with local news stories that show an increase in individuals seeking overdose prevention advice.

**Fig. 4:**
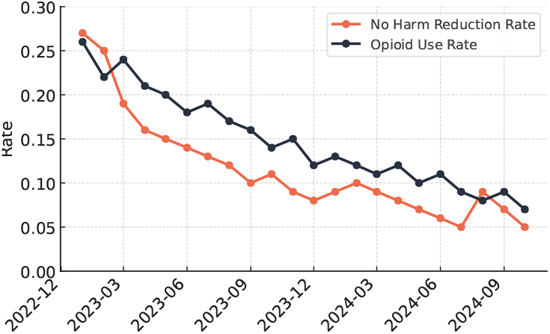
Trends in ‘No Harm Reduction’ mentions and ‘Opioid Use’ mentions over time. The downward trend in no-harm references implies broader awareness of safer practices, while self-reported opioid use shows moderate fluctuation rather than a sharp decline.

In contrast, the rate of opioid use remains relatively stable. Early commentary often included personal accounts of ongoing use, with some users attributing a sense of security to the absence of immediate arrest threats. Over time, however, the rate of explicit use disclosures saw mild oscillations rather than a clear upward or downward trend. Given that decriminalization alone may not solve underlying issues such as addiction severity or lack of mental health support, it is plausible that many who use opioids did not drastically alter their behavior. Instead, the major change is in how they discuss these matters publicly, shifting from coded or hidden references to more open acknowledgment of use combined with safer consumption tips.

From a technical poin of view, the zero-shot classification pipeline allowed us to parse implicit references to harm reduction or continued usage that did not always contain direct terms like ‘safe use’ or ‘opioid’. By referencing context (e.g., ‘I carry Narcan now’, ‘I’ve tried to cut back’), Mistral captured behaviors that simpler keyword-based approaches would likely miss. These findings confirm that while decriminalization may inspire people to explore safer practices, it does not, on its own, create uniform shifts in opioid consumption. Long-term trends likely require broader treatment availability, sustained harm reduction efforts, and ongoing public engagement.

### C. Limitations

Several limitations apply to this study. First, our reliance on zero-shot prompts with Mistral may miss certain domain-specific slang or subtle references if the context is too indirect. Although the model handles a wide range of linguistic forms, some user posts may involve novel expressions that exceed its pretrained knowledge. Second, the focus on Reddit discussions may not represent the entire population affected by opioid use or decriminalization. Individuals less familiar with online platforms or those unwilling to post in public forums may hold different attitudes. Third, our data captures a specific timeframe around the policy change and does not cover long-term effects on opioid use or public health. Post-policy developments, such as changes in treatment services, shifting law enforcement approaches, or broader community dynamics, may play a major role outside the observation window. Fourth, we only used Mistral for the classification tasks. Other LLMs might yield different outcomes, especially if they have different training data or attention mechanisms. These factors limit the direct interpretation of our findings as a complete picture of public sentiment and behavior in BC. The self-reported nature of online posts and their potential bias toward specific user groups should be noted when drawing conclusions about decriminalization’s overall impact.

## IV. Conclusion and Implications

This study examined Reddit discussions on opioid decriminalization in British Columbia to understand shifts in public sentiment (*S*(*c*_*i*_)) and self-reported behaviors (*B*(*c*_*i*_)). By leveraging Mistral’s attention-based architecture, we analyzed more than 22,000 comments to identify patterns in how people responded to the policy over time. We noted an initial burst of positive sentiment immediately following decriminalization, suggesting that early adopters of the discussion were optimistic about reduced stigma and improved access to harm reduction. As the discourse expanded, negative sentiment regained prominence, indicating skepticism about the policy’s long-term feasibility and potential unintended consequences. Self-reported behaviors related to opioid use and harm reduction showed parallel early engagement that later receded, likely reflecting a shift from curiosity and novelty to a more routine or critical view of the policy.

Although our zero-shot method efficiently captured evolving trends, the time-bound nature of the data limits insights into enduring policy outcomes. Future studies may integrate social media text with formal health records or epidemiological indicators to validate whether initial changes in sentiment align with real-world outcomes such as reduced overdose rates or increased treatment engagement. Additional work could involve fine-tuning LLMs on specialized corpora, enhancing their ability to detect complex or implicit references to opioid consumption and mental health. Researchers may also investigate cross-platform dynamics by comparing Reddit data with tweets, news articles, or official health advisories to obtain a more comprehensive picture of public attitudes. Combining advanced natural language processing with robust longitudinal datasets promises to yield deeper insights into how decriminalization policies influence both the discourse and the practical realities of opioid harm reduction.

## Data Availability

All data produced in the present study are available upon reasonable request to the authors

## ACKNOWLEDGMENT

This research is supported by the Artificial Intelligence for Public Health (AI4PH) Trainee Award funded by the Canadian Institutes of Health Research (CIHR), and the Natural Sciences and Engineering Research Council of Canada through the Canada Research Chairs program.

